# Percentile-Based Fetal Growth Velocity as a Predictor of Adverse Neonatal Outcomes in Fetal Growth Restriction and Small-for-Gestational-Age Pregnancies

**DOI:** 10.64898/2026.07.29.26359259

**Authors:** Joshua S. Brunton, Mohammad A. Salameh, Megan Branda, Ramila Mehta, Raymond C Stetson, Mauro Schenone, Kylie Cooper, Alyssa Larish, Regan N. Theiler

## Abstract

**Background:** Pregnancies complicated by fetal growth restriction are at increased risk of fetal demise and adverse neonatal outcomes. Distinguishing growth-restricted fetuses from constitutionally small ones remains challenging. Given the variability in current diagnostic criteria and the importance of identifying at-risk fetuses, fetal growth velocity has emerged as a predictor of adverse neonatal outcomes.

**Objective:** To evaluate whether percentile-based fetal growth velocity—defined as change in estimated fetal weight percentile per week—predicts adverse neonatal outcomes in pregnancies affected by fetal growth restriction or small-for-gestational-age neonates. The primary aim was to determine the relationship between growth velocity and a composite of adverse neonatal outcomes.

**Study Design:** This was a retrospective cohort study of pregnant patients 18–45 years old who delivered between August 2017 to December 2022 in a single healthcare system. Patients were excluded who had fewer than 2 ultrasounds after 16 weeks gestation, genetic or anatomic abnormalities, or a multiple gestation.

**Results:** 300 patients met all inclusion criteria, and most patients (n=199) delivered at the tertiary care center. Three had an intrauterine fetal demise at a mean gestational age of 35w3d. 74 neonates were admitted to the NICU with a mean length of stay of 8.5 days; 29 required respiratory support. No neonatal deaths occurred. In fetuses with initial estimated fetal weight <3rd percentile (n=33), the probability of composite outcome was increased (50%, 95% CI 44.8–55.2) compared to those >50th percentile (5%, 95% CI 3.7–6.6). In the 3–<10th and 10–50th percentile subgroups with decelerated growth, composite outcome rates were also increased (56.2% and 44.1%) compared to those with neutral or increased growth velocity.

**Conclusion:** Percentile-based fetal growth velocity is a simple calculation that correlates with adverse neonatal outcomes regardless of initial estimated fetal weight. As fetal growth velocity decreased, our cohort saw increased rates of adverse outcomes. Change in EFW percentile normalizes for gestational age and allows ease of clinical interpretation. Decelerated growth identified fetuses at highest risk, suggesting growth velocity as a useful metric in routine surveillance.

**Key Points:** **1.** Percentile-based fetal growth velocity correlates with adverse neonatal outcomes regardless of initial fetal size, including in fetuses with normal baseline estimated fetal weight. **2.** Percentile-based growth velocity is a clinically intuitive metric that normalizes for gestational age and may serve as a practical tool for risk stratification and surveillance in pregnancies affected by fetal growth restriction or at risk for small-for-gestational-age neonates.

## Introduction

Fetal growth restriction (FGR) is a major contributor to perinatal mortality and morbidity (Gregory et al. 2024; Resnik 2002; Kamphof et al. 2022). By identifying growth restricted fetuses during prenatal care, obstetric providers can implement surveillance to monitor for fetal compromise, improving outcomes (Martins et al. 2020; Hecher et al. 2001). Current definitions of FGR vary: the American College of Obstetricians and Gynecologists and Society for Maternal-Fetal Medicine define FGR as estimated fetal weight (EFW) or abdominal circumference <10th percentile (Martins et al. 2020; “Fetal Growth Restriction” 2021), while the International Society of Ultrasound in Obstetrics and Gynecology incorporates the Delphi consensus definition, utilizing biometric, Doppler, and growth trajectory parameters (Salomon et al. 2019).

The use of inconsistent diagnostic criteria reflects the challenges of distinguishing constitutionally small fetuses from those that are pathologically growth restricted—with the latter being at increased risk for stillbirth and adverse neonatal outcomes. Neonatal morbidity is higher among small-for-gestational age (SGA) neonates who are not diagnosed prenatally; however, detection rates for FGR remain low (Chauhan et al. 2014). One emerging metric to aid in detection is fetal growth velocity, or the change in fetal growth over time, which has been postulated as an additional approach to identification of fetuses experiencing fetal growth abnormalities (Stampalija et al. 2023; Bommarito et al. 2023; Ohuma et al. 2021; Sovio et al. 2015; Romero and Deter 2015; Jamieson-Grigg et al. 2025; Hiersch and Melamed 2018; D’Alberti et al. 2026). By categorizing pregnancies according to growth velocity, clinicians may better discern whether observed growth patterns are constitutionally determined or pathologic in origin (Grantz et al. 2018). Prior studies have calculated velocity in grams per week, a method that requires gestational age adjustment and is not easily translated into clinical practice (Stampalija et al. 2023). Percentile-based growth velocity, defined as change in EFW percentile per week, normalizes for gestational age and provides a clinically intuitive metric, yet remains understudied (Unterscheider et al. 2013; Gardosi et al. 2018).

We sought to determine the relationship between percentile-based growth velocity and adverse neonatal outcomes in a cohort of FGR and SGA affected pregnancies. Our hypothesis was that decelerated growth velocity would be associated with increased probability of adverse neonatal outcomes regardless of initial EFW percentile.

## Materials and Methods

### Study Cohort

A retrospective cohort was identified among pregnant subjects that delivered at the primary hospital and satellite health system hospitals between August 2017 to December 2022. Eligible deliveries were screened for the presence of any maternal or neonatal International Classification of Diseases, Ninth or Tenth Revision (ICD-9, ICD-10) codes for “fetal growth restriction”, “intrauterine growth restriction”, or “small for gestational age”. To best capture cases of fetal growth deceleration, subjects included those with normal antenatal EFW (10-90th percentile) who ultimately delivered SGA neonates. Subjects without legally-required research authorization, those with multiple gestations, known genetic syndromes (supplemental A), or known anatomic defects, and those with fewer than two ultrasounds performed after 16 weeks gestation were excluded (Figure 1). Eligible subjects had at least one baseline scan performed between 26w0d and 32w0d gestation, and one post-baseline scan between 32w0d and 36w6d gestation.

**Figure 1:**
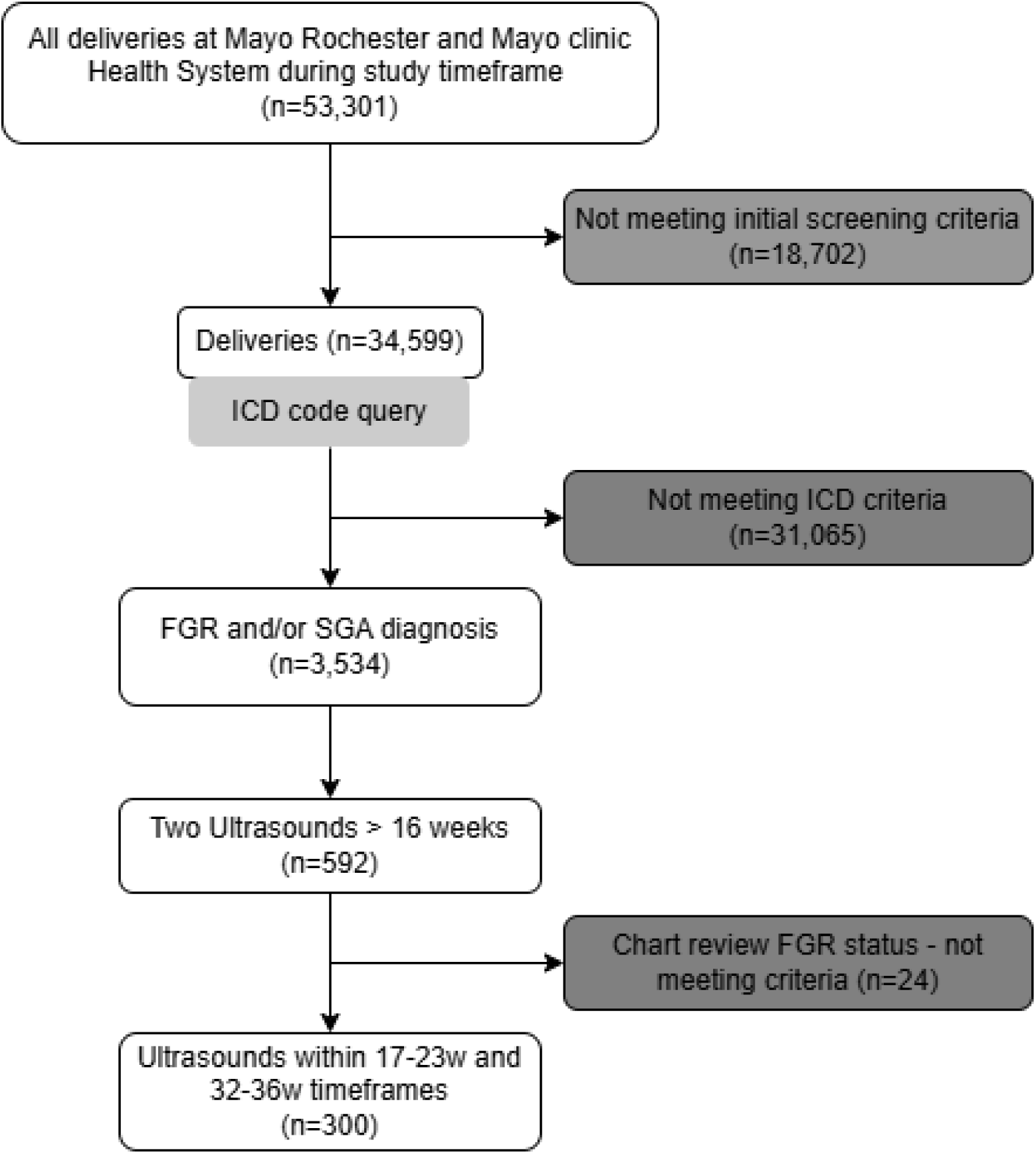
Participant selection flowchart.

### Data Collection

Patient information was collected using a validated registry of all deliveries within primary hospital and satellite health systems hospitals. Maternal and neonatal characteristics including demographic characteristics, laboratory values, delivery information, medications, and diagnoses were abstracted for all included patients. Ultrasound data was abstracted from the ClickView (Clickview Corporation, San Francisco CA) system for the primary hospital and manually abstracted from scanned ultrasound report documents for the satellite hospitals (JB and MS).

The ultrasound closest to the median timepoint for baseline and post-baseline points were chosen for analysis if a pregnancy had more than one ultrasound within the time-period. EFW was calculated using Hadlock formula 4: (Log_10_ (weight) = 1.3596 − 0.00386 × AC × FL + 0.0064 × HC + 0.00061 × BPD × AC + 0.0424 × AC + 0.174 × FL) (Hadlock et al. 1985). For study purposes, Hadlock growth percentiles were recalculated from ultrasound biometric measurements, regardless of whether an EFW percentile was formally reported. The baseline EFW was grouped into 4 clinically relevant categories: <3rd percentile, ≥3 to ≤10th percentile, >10 to ≤50th percentile, and >50th percentile.

### Growth Velocity Calculation

Growth velocity was calculated as the difference in EFW percentile between baseline and post-baseline ultrasounds divided by the difference in gestational age in weeks. The 26 0/7–31 6/7 week (baseline) and 32 0/7–36 6/7 week (post-baseline) intervals were selected to align with the timeframe in which most at risk-pregnancies undergo routine third-trimester growth surveillance in our institution. Because ultrasound timing varied in clinical practice, the widest reasonable windows were chosen to maximize sample size while preserving temporal separation for velocity estimation. Percentile-based velocity was chosen because it normalizes for gestational age, avoids the non-linear acceleration in grams/week expected in late gestation, and provides a clinically intuitive system of evaluation. We categorized velocity as accelerated (>+1 percentile/week), neutral (–1 to +1 percentile/week), or decelerated (<–1 percentile/week).

### Outcomes

The primary outcome is a composite of neonatal morbidity and severe adverse perinatal events. Neonatal morbidity was defined as any of: neonatal intensive care (NICU) admission, 5-minute APGAR score <7, intraventricular hemorrhage (IVH), necrotizing enterocolitis (NEC), or metabolic acidosis (umbilical artery pH 7.01–7.15 or base deficit 10–15.9 mmol/L). Severe adverse perinatal events were defined as stillbirth, neonatal death, hypoxic ischemic encephalopathy (HIE), need for mechanical ventilation, or severe metabolic acidosis (umbilical artery pH ≤7 or base deficit of ≥16 mmol/L) (Shankaran et al. 2005). As secondary outcomes, neonatal morbidity and severe adverse perinatal events were analyzed separately.

### Statistical Methods

Descriptive statistics are presented as means with 95% confidence intervals for continuous variables and as counts with percentages for categorical variables. Comparisons across baseline EFW categories for the neonatal outcomes were performed using the Fisher’s exact test.

Logistic regression was used to model the primary composite outcome as well as the secondary neonatal morbidity outcome. A univariate and multivariable model for each outcome was conducted. The multivariable model adjusted for fetal growth velocity (accelerated, decelerated and neutral), baseline EFW categories, pre-pregnancy maternal body mass index (BMI), gestational and pre-gestational hypertensive disorders, and GDM. For severe adverse perinatal events the model adjusted for velocity, pregravid BMI and pre-gestational hypertensive disorders only due to small event size. Odds ratios and 95% confidence intervals are reported. Assumptions of the models were verified, with the primary outcome model having a Hosmer-Lemeshow test (p = 0.47), AUC (0.74) indicated good fit and discrimination, and no influential observations (Cook’s distance > 0.5) noted within any models. Predicted probabilities of the composite outcome by fetal growth velocity were visualized in a scatter plot, stratified by baseline EFW categories, with a LOESS curve to smooth the trends. All statistical tests were two-sided, with significance set at α = 0.05. Analyses were conducted using SAS v9.4 (SAS Institute Inc., Cary, NC, USA).

## Results

### Pregnancy Outcomes

From August 2017 to December 2022, 592 patients were identified meeting criteria for inclusion (Figure 1). Out of these 592 patients, 300 patients had serial ultrasounds between the two study timepoints of 26–32 weeks and 32–36 weeks and were included in the growth velocity analysis. Baseline demographics, pregnancy outcomes, and ultrasound characteristics are summarized in Table 1. The mean gestation age of liveborn neonates was 37w6d (StdDev, ±1.75), and for those with known FGR, the mean length of time with a diagnosis of FGR was 4.4 weeks (StdDev, ±4.8). The majority (83.0%, n=249) of subjects delivered at term (≥37 weeks). Three subjects were diagnosed with an intrauterine fetal demise (IUFD), which was diagnosed at 34w3d (decreased growth velocity, FGR diagnosis), 35w3d (decreased growth velocity, FGR diagnosis), and 36w3d (unclear time since demise, pathology <3rd% for gestational age). The cesarean rate among all patients including planned cesareans was 33% (99) and about half (161, 49.7%) of the subjects underwent induction of labor.

**Table 1:** Baseline Characteristics.

|  | Baseline EFW |  |  |  |  |
| --- | --- | --- | --- | --- | --- |
|  | <=3% | 3-10% | 10-50% | >50% | Overall |
| **Sample size (n)** | 33 | 76 | 169 | 22 | 300 |
| **Maternal Demographics** |  |  |  |  |  |
| Age (years): Mean (95% CI) | 28.2 (26.2, 30.1) | 29.2 (27.9, 30.5) | 30.3 (29.4, 31.1) | 30.6 (28.4, 32.9) | 29.8 (29.2, 30.4) |
| Pre-pregnancy BMI (kg/m<sup>2</sup>): Mean (95% CI) | 26.3 (23.3, 29.4) | 26.3 (24.7, 27.8) | 27.3 (26.2, 28.4) | 29.2 (24.9, 33.5) | 27.0 (26.2, 27.9) |
| Missing | 1 | 1 | 9 | 2 | 13 |
| Ethnicity, n (%) |  |  |  |  |  |
| Hispanic or Latino | 1 (3.0%) | 4 (5.3%) | 13 (7.7%) | 2 (9.1%) | 20 (6.7%) |
| Not Hispanic or Latino | 32 (97.0%) | 72 (94.7%) | 155 (92.3%) | 20 (90.9%) | 279 (93.3%) |
| Missing | 0 | 0 | 1 | 0 | 1 |
| Smoking Current/Former, n (%) | 6 (19.4%) | 7 (13.0%) | 20 (14.3%) | 2 (10.0%) | 35 (14.3%) |
| Missing | 2 | 22 | 29 | 2 | 55 |
| Substance Current/Former, n (%) | 3 (9.7%) | 2 (3.7%) | 6 (4.3%) | 0 (0.0%) | 11 (4.5%) |
| Missing | 2 | 22 | 29 | 2 | 55 |
| **Pregnancy Complications** |  |  |  |  |  |
| Chronic Hypertension, n (%) | 4 (12.1%) | 3 (3.9%) | 20 (11.8%) | 2 (9.1%) | 29 (9.7%) |
| Hypertensive disorder of pregnancy, n (%) | 13 (39.4%) | 15 (19.7%) | 31 (18.3%) | 5 (22.7%) | 64 (21.3%) |
| Gestational hypertension Or pre-eclampsia without severe features, n (%) | 12 (36.4%) | 12 (15.8%) | 25 (14.8%) | 5 (22.7%) | 54 (18.0%) |
| Pre-eclampsia with severe features or HELLP, n (%) | 5 (15.2%) | 6 (7.9%) | 10 (5.9%) | 2 (9.1%) | 23 (7.7%) |
| Anti hypertensive Medications, n (%) | 6 (18.2%) | 7 (9.2%) | 25 (14.8%) | 5 (22.7%) | 43 (14.3%) |
| Gestational Diabetes, n (%) | 6 (18.2%) | 12 (15.8%) | 31 (18.3%) | 8 (36.4%) | 57 (19.0%) |
| Pre-Gestational Diabetes, n (%) | 2 (6.1%) | 3 (3.9%) | 4 (2.4%) | 2 (9.1%) | 11 (3.7%) |
| **Ultrasound Characteristics** |  |  |  |  |  |
| GA at baseline US (weeks): Mean (95% CI) | 28.8 (28.1, 29.5) | 28.7 (28.2, 29.2) | 28.7 (28.4, 29.0) | 28.3 (27.6, 29.1) | 28.7 (28.5, 28.9) |
| GA at follow-up US (weeks): Mean (95% CI) | 34.6 (34.1, 35.0) | 35.0 (34.7, 35.2) | 34.7 (34.5, 34.9) | 34.2 (33.5, 34.8) | 34.7 (34.6, 34.9) |
| Time interval (weeks): Mean (95% CI) | 5.8 (5.0, 6.6) | 6.3 (5.7, 6.8) | 6.0 (5.7, 6.3) | 5.9 (5.0, 6.7) | 6.0 (5.8, 6.3) |
| Growth trend, n (%) |  |  |  |  |  |
| Increased | 7 (21.2%) | 30 (39.5%) | 31 (18.3%) | 1 (4.5%) | 69 (23.0%) |
|  | <=3% | 3-10% | 10-50% | >50% | Overall |
| Decreased | 0 (0.0%) | 3 (3.9%) | 91 (53.8%) | 20 (90.9%) | 114 (38.0%) |
| Neutral | 26 (78.8%) | 43 (56.6%) | 47 (27.8%) | 1 (4.5%) | 117 (39.0%) |
| SGA/FGR Status, n (%) |  |  |  |  |  |
| FGR | 10 (30.3%) | 27 (35.5%) | 43 (25.4%) | 5 (22.7%) | 85 (28.3%) |
| SGA | 0 (0.0%) | 6 (7.9%) | 67 (39.6%) | 15 (68.2%) | 88 (29.3%) |
| SGA/FGR | 23 (69.7%) | 43 (56.6%) | 59 (34.9%) | 2 (9.1%) | 127 (42.3%) |
| Time with FGR Diagnosis (weeks):<br>Mean (95% CI) | 6.8 (5.1, 8.4) | 5.7 (4.5, 6.9) | 2.7 (1.9, 3.5) | 4.2 (0, 11.3) | 4.4 (3.7, 5.0) |
| **Delivery Characteristics** |  |  |  |  |  |
| GA at Delivery (Weeks): Mean<br>(95% CI) | 36.5 (35.8, 37.1) | 38.0 (37.6, 38.4) | 37.9 (37.6, 38.4) | 38.5 (38.0, 39.0) | 37.8 (37.6, 38.0) |
| Preterm Premature Rupture of<br>Membranes, n (%) | 2 (6.1%) | 0 (0.0%) | 8 (4.7%) | 0 (0.0%) | 10 (3.3%) |
| Meconium Baby or Mom Dx, n<br>(%) | 1 (3.0%) | 4 (5.3%) | 15 (8.9%) | 1 (4.5%) | 21 (7.0%) |
| Chorioamnionitis, n (%) | 0 (0.0%) | 4 (5.3%) | 6 (3.6%) | 1 (4.5%) | 11 (3.7%) |
| Placenta Abruption, n (%) | 0 (0.0%) | 1 (1.3%) | 1 (0.6%) | 0 (0.0%) | 2 (0.7%) |
| Delivery Method, n (%) |  |  |  |  |  |
| Cesarean Section | 12 (36.4%) | 21 (27.6%) | 59 (34.9%) | 7 (31.8%) | 99 (33.0%) |
| VBAC, Spontaneous | 1 (3.0%) | 1 (1.3%) | 3 (1.8%) | 0 (0.0%) | 5 (1.7%) |
| Vaginal, Forceps | 0 (0.0%) | 1 (1.3%) | 2 (1.2%) | 1 (4.5%) | 4 (1.3%) |
| Vaginal, Spontaneous | 20 (60.6%) | 52 (68.4%) | 101 (59.8%) | 13 (59.1%) | 186 (62.0%) |
| Vaginal, Vacuum | 0 (0.0%) | 1 (1.3%) | 4 (2.4%) | 1 (4.5%) | 6 (2.0%) |
| Birth Weight (grams):<br>Mean (95% CI) | 2140.4 (1960, 2321) | 2574.9 (2482, 2667) | 2573.6 (2507, 2640) | 2839.1 (2668, 3011) | 2545.7 (2493, 2598) |
| Male sex, n (%) | 11 (33.3%) | 19 (25.0%) | 63 (37.3%) | 10 (45.5%) | 103 (34.3%) |

### Neonatal Outcomes

Most neonates were female (65.7%, 197) with a mean birthweight of 2545.7 grams (range 1150–4260, ±461.5) and mean APGAR scores at 1 and 5 minutes of 7.7 (range 1–9, StdDev ±1.43) and 8.7 (range 3–10, StdDev ±0.78), respectively. 74 of the 300 (24.7%) were admitted to the neonatal intensive care unit (NICU) and 39.1% (29/74) of those were admitted for respiratory indications. There was a significant difference in the gestational age at time of delivery between groups, with 39.4% of the <3rd percentile group being delivered in the 32w0d–36w6d timeframe compared to 13.6%, 16%, and 4.6% in those with baseline EFW 3–10%ile, 10–50%ile, and >50%ile respectively (p-value = 0.002). No neonatal deaths occurred, and NICU length of stay averaged 8.5 days (range 0.5–35.5) following admission with a mean neonatal discharge weight of 2520.1 grams (range 1490–4135). The remaining neonatal outcomes can be viewed in Table 2. Supplemental tables 1–4 show demographics and outcomes for those with FGR, excluding cases with only SGA.

**Table 2:**
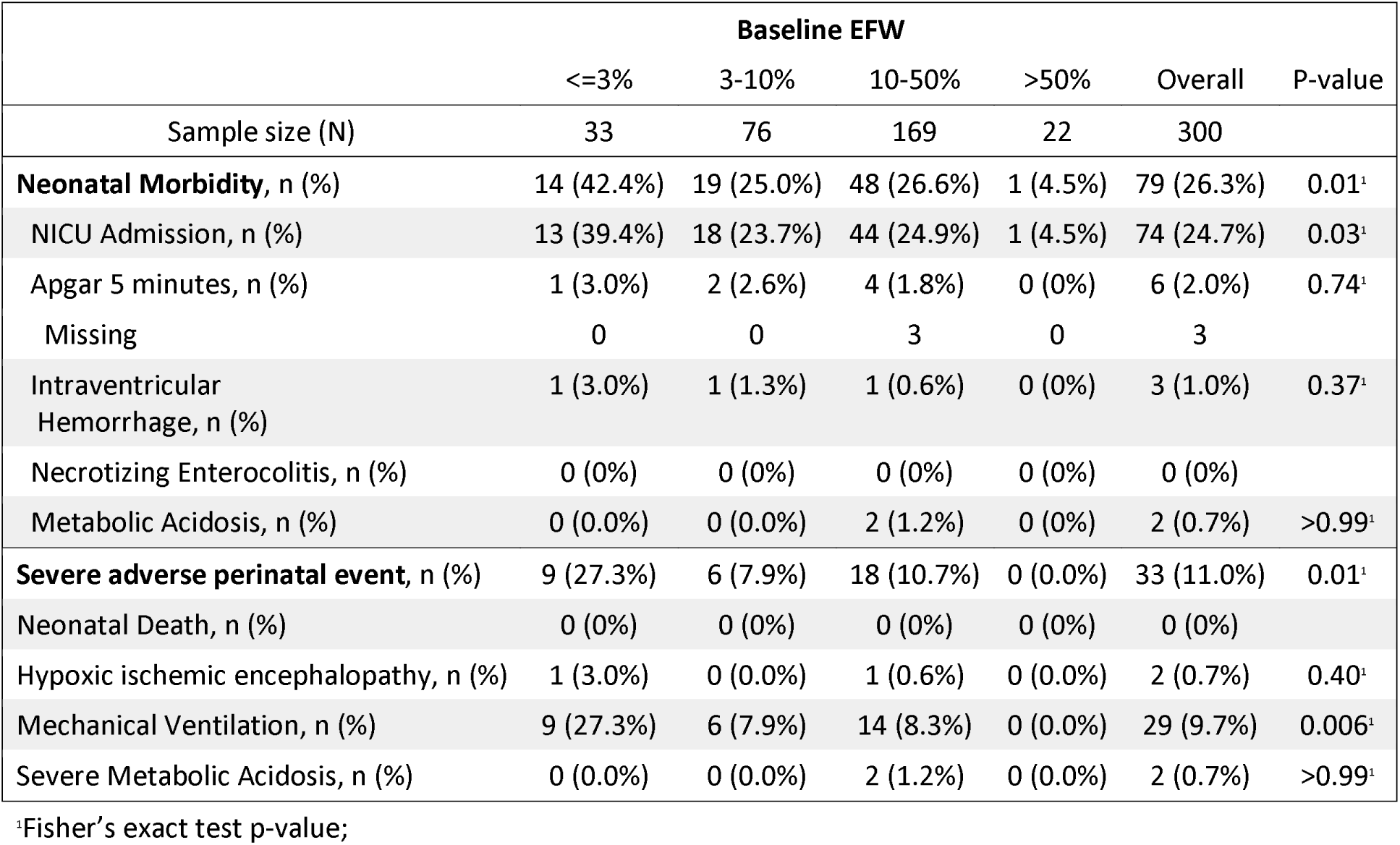
Neonatal outcomes.

### Growth Velocity

Within all baseline fetal growth categories, growth velocity correlated with the composite adverse outcomes, with decelerated growth associated with increased rates of the composite outcome (Table 3, Figure 2). For the pooled analysis of all baseline groups, the probability of the composite outcome increased as growth velocity decreased, adjusted OR 3.57, 95% CI 1.60–7.96 (Table 3). Within the baseline EFW <3rd percentile patients the highest incidence of the composite adverse neonatal outcome occurred (50%, 95% CI 44.8–55.2), occurring in 55.3% (95% CI 50.6–60.0) of patients with neutral growth velocity and in 31.0% (95% CI 26.6–35.4) of those with accelerated growth. Among the 3–10%ile and 10–50%ile EFW baseline subgroups with decelerated growth, the probability of the composite outcome occurred at a rate of 56.2% (95% CI 51.6–60.8) and 44.1% (95% CI 42.0–46.1), which was significantly more frequent than in those with neutral or accelerated growth (Table 4). As pictured in Figure 2, the composite outcome correlated with both baseline EFW percentile and subsequent fetal growth velocity, with decelerated fetal growth correlating with more adverse neonatal outcomes across all baseline EFW categories.

**Figure 2:**
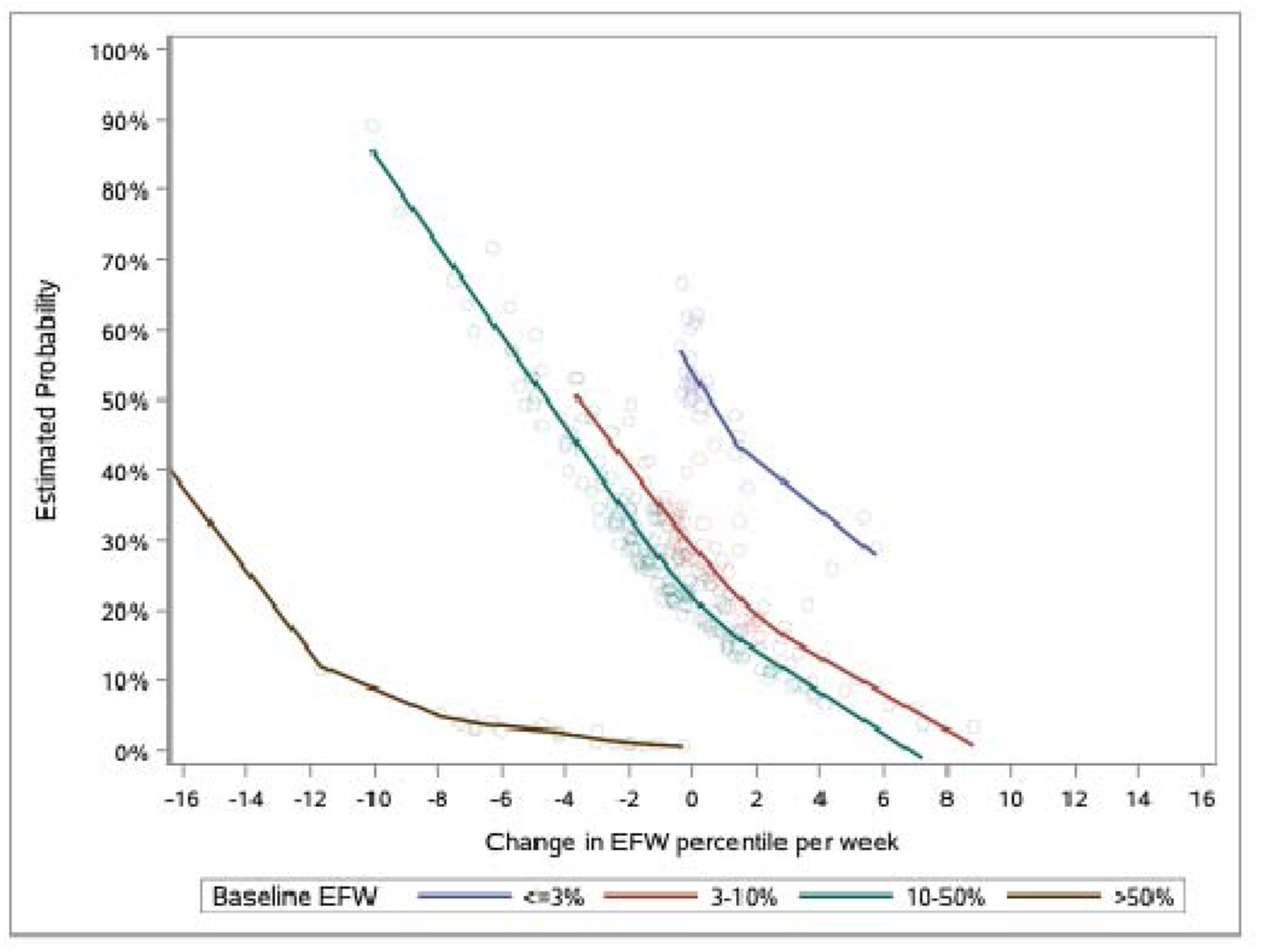
Probability of composite outcome occurring based on fetal growth velocity. Logistic model adjusted by change in EFW percentile per week, baseline EFW and pregravid BMI. The predicted probabilities of the composite outcome with a LOESS curve to smooth the trends.

**Table 3:** Primary and secondary outcomes by growth velocity.

| Primary Outcome | Velocity Group | Events/N | Unadjusted OR (95% CI) | Adjusted OR (95% CI)<sup>3</sup> |
| --- | --- | --- | --- | --- |
| Composite of Neonatal morbidity & Severe Adverse Perinatal Event | Accelerated Growth | 10/69 | 0.40 (0.18, 0.87) | 0.47 (0.20, 1.10) |
|  | Neutral Growth | 35/117 | Ref | Ref |
|  | Decelerated Growth | 43/114 | 1.42 (0.82, 2.45) | 3.57 (1.60, 7.96) |
| **Secondary Outcomes** |  |  |  |  |
| Neonatal Morbidity<sup>1</sup> | Accelerated Growth | 10/69 | 0.45 (0.21, 0.99) | 0.54 (0.23, 1.26) |
|  | Neutral Growth | 32/117 | Ref | Ref |
|  | Decelerated Growth | 37/114 | 1.28 (0.73, 2.25) | 2.85 (1.26, 6.46) |
| Severe Adverse Perinatal Event | Accelerated Growth | 6/69 | 0.76 (0.28, 2.11) | 0.82 (0.27, 2.52) |
|  | Neutral Growth | 13/117 | Ref | Ref |
|  | Decelerated Growth | 14/114 | 1.12 (0.50, 2.50) | 0.99 (0.42, 2.33) |

<sup>1</sup>Model adjusted by velocity, EFW baseline, pregravid BMI, gestational diabetes, hypertensive disorder of pregnancy.
<sup>2</sup>Model adjusted by velocity, pregravid BMI and hypertensive disorder of pregnancy.
<sup>3</sup>Observations missing pre-gravid BMI N = 13 are omitted from analysis.

**Table 4:**
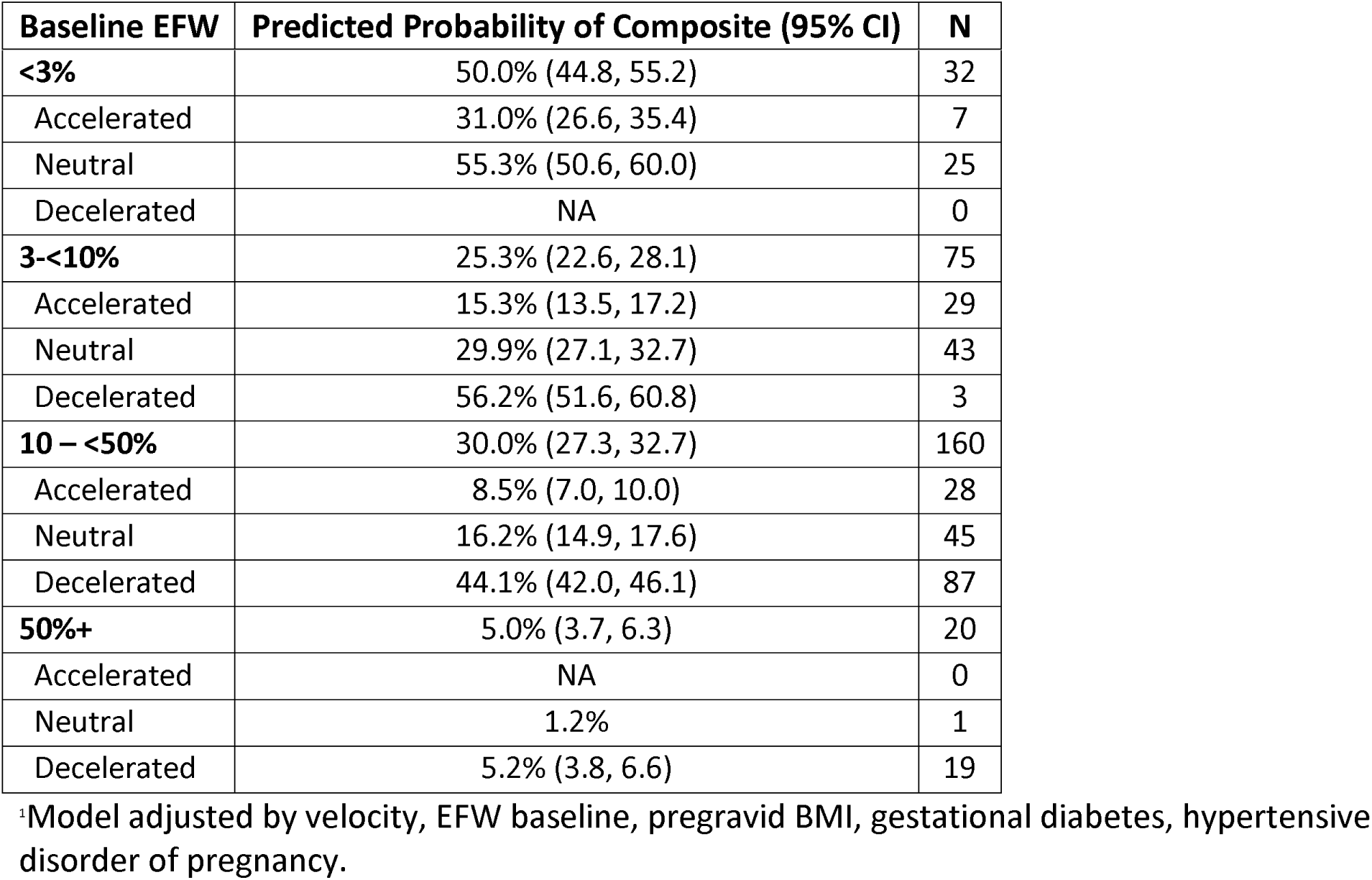
Average predicted probability of neonatal composite outcome by baseline EFW and growth velocity.^1^.

| Baseline EFW | Predicted Probability of Composite (95% CI) | N |
| --- | --- | --- |
| **<3%** | 50.0% (44.8, 55.2) | 32 |
| Accelerated | 31.0% (26.6, 35.4) | 7 |
| Neutral | 55.3% (50.6, 60.0) | 25 |
| Decelerated | NA | 0 |
| **3-<10%** | 25.3% (22.6, 28.1) | 75 |
| Accelerated | 15.3% (13.5, 17.2) | 29 |
| Neutral | 29.9% (27.1, 32.7) | 43 |
| Decelerated | 56.2% (51.6, 60.8) | 3 |
| **10 – <50%** | 30.0% (27.3, 32.7) | 160 |
| Accelerated | 8.5% (7.0, 10.0) | 28 |
| Neutral | 16.2% (14.9, 17.6) | 45 |
| Decelerated | 44.1% (42.0, 46.1) | 87 |
| **50%+** | 5.0% (3.7, 6.3) | 20 |
| Accelerated | NA | 0 |
| Neutral | 1.2% | 1 |
| Decelerated | 5.2% (3.8, 6.6) | 19 |

<sup>‡</sup>Model adjusted by velocity, EFW baseline, pregravid BMI, gestational diabetes, hypertensive disorder of pregnancy.

## Discussion

Our study calculated fetal growth velocities among a population with FGR or SGA neonates, demonstrating an increasing probability of an adverse neonatal outcome with decreasing growth velocity. This finding is also consistent with prior studies (Stampalija et al. 2023; Jamieson-Grigg et al. 2025; Grantz et al. 2018), in which growth velocity was assessed in grams per week, showed an increase in adverse perinatal outcomes with decreased growth velocity. Our study differs from many other studies by utilizing the more clinically intuitive velocity measure of change in growth percentile over time. This approach normalized for the non-linearity of the standard fetal growth curve, making it easier to apply across gestation compared to growth velocity measured in grams per week (Grantz et al. 2018).

A paucity of existing literature examines fetal growth change in percentiles, and the available literature derives largely from a consensus of experts and one study examining mid trimester growth velocity in appropriate-for-gestational-age infants (Gordijn et al. 2016; Kennedy et al. 2020). Similarly to that study, we included SGA neonates without prenatal diagnoses of FGR and retrospectively assessed fetal growth velocity. Our findings suggest that even for the fetus with normal baseline EFW (10th– 90th percentile), decreasing EFW percentile between sequential ultrasounds correlates with adverse neonatal outcomes. It reinforces the prognostic value of not only low initial EFW, but also deceleration in growth, supporting the potential integration of growth velocity into routine risk stratification and delivery planning in high-risk pregnancies.

This study has several strengths. EFW was recalculated using the Hadlock formula from primary ultrasound measurements, minimizing variability from differing equipment calculations across sites. The multicenter design (8 sites) increased sample size and reduced site-specific bias. Inclusion of SGA neonates regardless of antenatal FGR diagnosis helped mitigate the well-documented challenges of imperfect sonographic detection of growth-restricted fetuses (Fadigas et al. 2015; Chauhan et al. 2014). Several limitations warrant consideration. First, sonographic EFW estimation is inherently imprecise (Ma et al. 2023; Chien 2000), and measurement variability may influence percentile-based velocity calculations; however, reliance on ultrasound data enhances real-world applicability. Second, this retrospective analysis did not employ standardized gestational age timepoints for velocity assessment, which may introduce heterogeneity but mirrors routine clinical practice (Stampalija et al. 2023; Ohuma et al. 2021). Third, inclusion of postnatally identified SGA neonates limits prospective application during pregnancy, and those diagnosed prenatally with FGR were likely managed differently, impacting delivery outcomes. Finally, the absence of a control group without suspected growth restriction precluded estimation of normative growth velocities and baseline complication rates.

Future work should focus on prospective validation of our findings, in addition to comparison of the predictive value of growth velocity analysis to that of umbilical artery Doppler studies in FGR fetuses. Additionally, stable growth velocity may serve to distinguish truly growth restricted fetuses from constitutionally small ones, and this approach requires further validation. Finally, our data hints that negative growth velocity, even in non-FGR fetuses, may be an indicator of increased risk of adverse events; this hypothesis should be separately validated among prospective cohorts of initially normal sized fetuses.

## Conclusion

The diagnosis of FGR is filled with imprecision as obstetricians try to distinguish pathologically growth restricted fetuses from constitutionally small ones. Percentile-based fetal growth velocity represents a simple, clinically interpretable tool that may help identify growth-restricted fetuses at elevated risk of adverse neonatal outcomes. In this retrospective cohort, fetuses with accelerated growth had the most favorable outcomes, whereas those with neutral or decelerated growth had higher rates of morbidity—particularly among those with low initial EFW percentiles. Although promising, this approach requires prospective validation, incorporation of standardized ultrasound timing, and comparison with alternative velocity metrics before routine clinical implementation.

## Data Availability

All data produced in the present study are available upon reasonable request to the authors

## SUPPLEMENTAL TABLES AND FIGURES

**Table S1:** Baseline Characteristics - FGR diagnosed fetuses only.

|  | Baseline EFW |  |  |  | Overall |
| --- | --- | --- | --- | --- | --- |
|  | <=3% | 3-10% | 10-50% | >50% |  |
| **Sample size** | 33 | 70 | 102 | 7 | 212 |
| **Maternal Demographics** |  |  |  |  |  |
| Age (years): Mean (95% CI) | 28.2 (26.2, 30.1) | 29.0 (27.6, 30.3) | 29.8 (28.8, 30.8) | 29.4 (24.6, 34.3) | 29.3 (28.5, 30.0) |
| Pre-pregnancy BMI: Mean (95% CI) | 26.3 (26.2, 30.1) | 26.3 (24.6, 27.9) | 26.9 (25.4, 28.4) | 27.8 (20.4, 35.2) | 26.6 (25.6, 27.6) |
| Missing | 1 | 1 | 8 | 1 | 11 |
| Ethnicity, n (%) |  |  |  |  |  |
| Hispanic or Latino | 1 (3.0%) | 4 (5.7%) | 9 (8.9%) | 0 (0%) | 14 (6.6) |
| Not Hispanic or Latino | 32 (97.0%) | 66 (94.3%) | 92 (91.1%) | 7 (100%) | 197 (93.4%) |
| Missing | 0 | 0 | 1 | 0 | 1 |
| Smoking Current/Former, n (%) | 6 (19.4%) | 6 (12.5%) | 9 (10.6%) | 1 (14.3%) | 22 (12.9%) |
| Missing | 2 | 22 | 17 | 0 | 41 |
| Substance Current/Former, n (%) | 3 (9.7%) | 2 (4.2%) | 3 (3.5%) | 0 (0.0%) | 8 (4.7%) |
| Missing | 2 | 22 | 17 | 0 | 41 |
| **Pregnancy Complications** |  |  |  |  |  |
| Chronic Hypertension, n (%) | 4 (12.1%) | 2 (2.9%) | 10 (9.8%) | 0 (0%) | 16 (7.5%) |
| Hypertensive disorder of pregnancy, n (%) | 13 (39.4%) | 15 (21.4%) | 19 (18.6%) | 2 (28.6%) | 49 (23.1%) |
| Gestational hypertension Or pre-eclampsia without severe features, n (%) | 12 (36.4%) | 12 (17.1%) | 14 (13.7%) | 2 (28.6%) | 49 (23.1%) |
| Pre-eclampsia with severe features or HELLP, n (%) | 5 (15.2%) | 6 (8.6%) | 8 (8.7%) | 0 (0%) | 19 (9.0%) |
| Antihypertensive Medications, n (%) | 6 (18.2%) | 7 (10.0%) | 19 (18.6%) | 1 (14.3%) | 33 (15.6%) |
| Gestational Diabetes, n (%) | 6 (18.2%) | 11 (15.7%) | 15 (14.7%) | 2 (28.6%) | 34 (16.0%) |
| Pre-Gestational Diabetes, n (%) | 2 (6.1%) | 3 (4.3%) | 2 (2.0%) | 1 (14.3%) | 8 (3.8%) |
| **Ultrasound Characteristics** |  |  |  |  |  |
| GA at baseline US (weeks): Mean (95% CI) | 28.8 (28.1, 29.5) | 28.7 (28.2, 29.2) | 28.7 (28.3, 29.0) | 28.7 (26.5, 30.9) | 28.7 (28.4, 28.9) |
| GA at follow-up US (weeks): Mean (95% CI) | 34.6 (34.1, 35.0) | 35.0 (34.7, 35.3) | 34.5 (34.3, 34.8) | 33.7 (32.1, 35.3) | 34.7 (34.5, 34.9) |
| Time interval US (weeks): Mean 95% CI) | 5.8 (5.0, 6.6) | 6.3 (5.7, 6.9) | 5.9 (5.5, 6.2) | 5.0 (3.2, 6.8) | 6.0 (5.7, 6.3) |
| Growth trend, n (%) |  |  |  |  |  |
| Increased | 7 (21.2%) | 24 (34.3%) | 14 (13.7%) | 0 (0%) | 45 (21.2%) |
| Decelerated | 0 (0.0%) | 3 (4.3%) | 62 (60.8%) | 7 (100%) | 72 (34.0%) |
| Neutral | 26 (78.8%) | 43 (61.4%) | 26 (25.5%) | 0 (0%) | 95 (44.8%) |
| SGA/FGR Status, n (%) |  |  |  |  |  |
| FGR | 10 (30.3%) | 27 (38.6%) | 43 (42.2%) | 5 (71.4%) | 85 (40.1%) |
| SGA/FGR | 23 (69.7%) | 43 (61.4%) | 59 (57.8%) | 2 (28.6%) | 127 (59.9%) |
| Time with FGR Diagnosis (weeks): Mean (95% CI) | 6.8 (5.1, 8.4) | 5.7 (4.5, 6.9) | 2.7 (1.9, 3.5) | 4.2 (0, 11.3) | 4.4 (3.7, 5.0) |
| **Delivery Characteristics** |  |  |  |  |  |
| GA at Delivery (Weeks): Mean (95% CI) | 36.5 (35.8, 37.1) | 37.9 (37.5, 38.3) | 37.5 (37.1, 37.9) | 38.7 (37.7, 39.7) | 37.5 (37.3, 37.8) |
| Preterm Premature Rupture of Membranes, n (%) | 2 (6.1%) | 0 (0.0%) | 6 (5.9%) | 0 (0.0%) | 8 (3.8%) |
| Meconium Baby or Mom Dx, n (%) | 1 (3.0%) | 4 (5.7%) | 5 (4.9%) | 0 (0.0%) | 10 (4.7%) |
| Chorioamnionitis, n (%) | 0 (0.0%) | 4 (5.7%) | 4 (3.9%) | 1 (14.3%) | 9 (4.2%) |
| Placenta Abruption, n (%) | 0 (0.0%) | 1 (1.4%) | 1 (1.0%) | 0 (0.0%) | 2 (0.9%) |
| Delivery Method, n (%) |  |  |  |  |  |
| C-Section | 12 (36.4%) | 20 (28.6%) | 43 (42.2%) | 2 (28.6%) | 77 (36.3%) |
| VBAC, Spontaneous | 1 (3.0%) | 1 (1.4%) | 1 (1.0%) | 0 (0.0%) | 3 (1.4%) |
| Vaginal, Forceps | 0 (0.0%) | 1 (1.4%) | 0 (0.0%) | 0 (0.0%) | 1 (0.5%) |
| Vaginal, Spontaneous | 20 (60.6%) | 47 (67.1%) | 54 (52.9%) | 5 (71.4%) | 126 (59.4%) |
| Vaginal, Vacuum | 0 (0.0%) | 1 (1.4%) | 4 (3.9%) | 0 (0.0%) | 5 (2.4%) |
| Birth Weight (grams): Mean (95% CI) | 2140.4 (1960, 2321) | 2567.0 (2467, 2667) | 2544.3 (2444, 2645) | 3141.4 (2650, 3633) | 2509 (2439, 2579) |
| Male sex, n(%) | 11 (33.3%) | 19 (27.1%) | 33 (32.4%) | 2 (28.6%) | 65 (30.7%) |

**Table S2:** Neonatal outcomes-FGR diagnosed fetuses only.

|  | Baseline EFW |  |  |  | Overall | P-value<sup>2</sup> |
| --- | --- | --- | --- | --- | --- | --- |
|  | <=3% | 3-10% | 10-50% | >50% |  |  |
| Sample size (N) | 33 | 70 | 102 | 7 | 212 |  |
| **Neonatal Morbidity, n (%)** | 14<br>(42.4%) | 18<br>(25.7%) | 34<br>(33.3%) | 0<br>(0.0%) | 66<br>(31.1%) | 0.10 |
| NICU Admission, n (%) | 13<br>(39.4%) | 17<br>(24.3%) | 33<br>(32.4%) | 0<br>(0.0%) | 63<br>(29.7%) | 0.13 |
| Apgar 5 minutes < 7<sup>1</sup>, n (%) | 1 (3.0%) | 2 (2.9%) | 1 (1.0%) | 0<br>(0.0%) | 4 (1.9%) | 0.56 |
| Intraventricular Hemorrhage, n (%) | 1 (3.0%) | 1 (1.4%) | 0 (0.0%) | 0<br>(0.0%) | 2 (0.9%) | 0.19 |
| Necrotizing Enterocolitis, n (%) | 0 (0%) | 0 (0%) | 0 (0%) | 0 (0%) | 0 (0%) |  |
| Metabolic Acidosis, n (%) | 0 (0.0%) | 0 (0.0%) | 2 (2.0%) | 0<br>(0.0%) | 2 (0.9%) | 0.19<sup>1</sup> |
| **Severe adverse perinatal event, n (%)** | 9 (27.3%) | 6 (8.6%) | 15<br>(14.7%) | 0<br>(0.0%) | 30<br>(14.2%) | 0.07<sup>1</sup> |
| Neonatal Death, n (%) | 0 (0%) | 0 (0%) | 0 (0%) | 0 (0%) | 0 (0%) |  |
| Hypoxic ischemic encephalopathy, n (%) | 1 (3.0%) | 0 (0.0%) | 1 (1.0%) | 0<br>(0.0%) | 2 (0.9%) | 0.45 |
| Mechanical Ventilation n (%) | 9 (27.3%) | 6 (8.6%) | 12<br>(11.8%) | 0<br>(0.0%) | 27<br>(12.7%) | 0.06 |
| Severe Metabolic Acidosis, n (%) | 0 (0.0%) | 0 (0.0%) | 2 (2.0%) | 0<br>(0.0%) | 2 (0.9%) | 0.68 |

1 – Missing Apgar score at 5 minutes for 2 patients in the Baseline EFW category of 10-50%
2 – Fisher's Exact p-value

**Table S3:** Primary and secondary outcomes by growth velocity - FGR diagnosed fetuses only.

| Primary Outcome | Velocity Group | Events/N | Unadjusted OR<br>(95% CI)<sup>1</sup> | Adjusted OR<br>(95% CI)<sup>2, 3</sup> |
| --- | --- | --- | --- | --- |
| **Composite of Neonatal morbidity & Severe Adverse Perinatal Event** | Accelerated Growth | 7/45 | 0.33 (0.13, 0.82) | 0.33 (0.12, 0.91) |
|  | Neutral Growth | 34/95 | Ref | Ref |
|  | Decelerated Growth | 33/65 | 1.85 (0.97, 3.52) | 2.64 (1.04, 6.73) |
| **Secondary Outcomes** |  |  |  |  |
| **Neonatal Morbidity** | Accelerated Growth | 7/45 | 0.38 (0.14, 0.95) | 0.38 (0.14, 1.06) |
|  | Neutral Growth | 31/95 | Ref | Ref |
|  | Decelerated Growth | 28/65 | 1.56 (0.81, 3.00) | 2.03 (0.78, 5.24) |
| **Severe Adverse Perinatal Event** | Accelerated Growth | 5/45 | 0.79 (0.26, 2.37) | 1.04 (0.28, 3.85) |
|  | Neutral Growth | 13/95 | Ref | Ref |
|  | Decelerated Growth | 12/65 | 1.43 (0.61, 3.37) | 1.72 (0.42, 7.00) |

<sup>1</sup>Observations removed from analysis with baseline EFW > 50% (N=7).
<sup>2</sup>Model adjusted by velocity, EFW baseline, pregravid BMI, gestational diabetes, hypertensive disorder of pregnancy.
<sup>3</sup>Observations missing pre-gravid BMI N = 11 are omitted from analysis as well as baseline EFW > 50% (N=7), total N=17 patients (N=1 for BMI and EFW > 50%).

**Table S4:** Average predicted probability of neonatal composite outcome by baseline EFW and growth velocity - FGR diagnosed fetuses only.^1^.

| Baseline EFW | Predicted Probability of Composite (95% CI) | N |
| --- | --- | --- |
| **<3%** | 50% (43.0, 57.0) | 32 |
| Accelerated | 25.7% (17.2, 34.1) | 7 |
| Neutral | 56.8% (50.3, 63.3) | 25 |
| Decelerated | NA | 0 |
| **3-<10%** | 26.1% (22.4, 29.8) | 69 |
| Accelerated | 12.6% (9.7, 15.4) | 23 |
| Neutral | 31.7% (27.5, 35.9) | 43 |
| Decelerated | 49.7% (39.7, 59.6) | 3 |
| **10 – <50%** | 39.4% (35.4, 43.3) | 94 |
| Accelerated | 11.9% (4.8, 19.1) | 11 |
| Neutral | 24.7% (21.4, 28.0) | 25 |
| Decelerated | 50.9% (47.3, 54.5) | 58 |

<sup>1</sup>Model adjusted by velocity, EFW baseline, pregravid BMI, gestational diabetes, hypertensive disorder of pregnancy.

**Figure S1:**
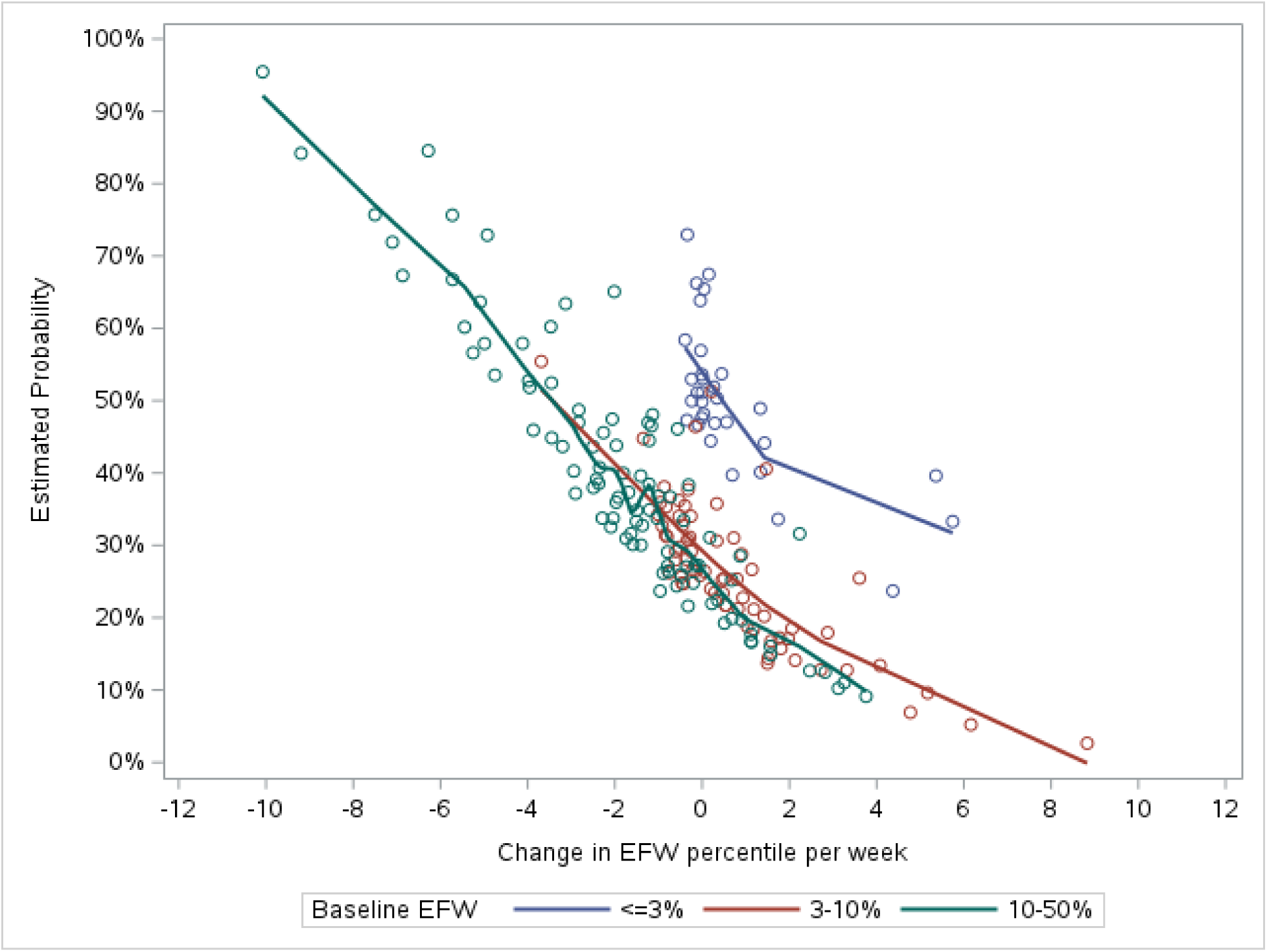
Probability of composite outcome occurring based on fetal growth velocity – FGR diagnosed fetuses only.

## Notes

### Competing Interest Statement

R.N.T. declares the following: Delfina Care Inc.--Medical Advisory Board, Sanofi--Medical Advisory Board, Beckman Coulter -- consultant. R.C.S. declares the following: Receipt of Moderna sponsored grant funding in support of retrospective population-based research on neonatal and pediatric infectious conditions. The remaining authors report no conflicts of interest.

### Author Declarations

Ethics committee/IRB of Mayo Clinic gave ethical approval for this work.

